# Combined vitrectomy, near-confluent endolaser, bevacizumab and cyclophotocoagulation for neovascular glaucoma

**DOI:** 10.1101/2020.01.19.20017889

**Authors:** P. Strzalkowski, A. Strzalkowska, W. Göbel, T. Ach, N.A. Loewen, J. Hillenkamp

## Abstract

**Purpose:** Evaluated the safety and efficacy of an integrative surgical approach to neovascular glaucoma (NVG).

**Methods:** Consecutive interventional case series of NVG with one-year follow-up. Eyes underwent pars plana vitrectomy, near-confluent panretinal photocoagulation, intravitreal bevacizumab, and transscleral cyclophotocoagulation. Phakic eyes underwent concomitant cataract surgery. Best-corrected visual acuity (BCVA, logMAR), intraocular pressure (IOP, mmHg), glaucoma medication score (GMS), visual analog pain scale (VAPS, 0-10) were recorded at baseline, and 1, 3, 6 and 12 months. Blind eyes were excluded.

**Results:** 83 eyes of 83 patients (53 male, 30 female, mean age 74.6±11.6 years) were included and 53 completed a one-year follow-up. NVG underlying conditions included retinal vein occlusion (n=41), proliferative diabetic retinopathy (n=25), central retinal artery occlusion (n=10), and ocular ischemic syndrome (n=6). Mean IOP decreased postoperatively from 46.0±10.3 mmHg to 14.2±8.9 mmHg (p<0.001), GMS from 4.8 to 1.8 (p<0.001) and VAPS from 6 to 0. BCVA was unchanged. All postoperative complications had resolved at 1 month postop. 26 eyes did not require additional surgical treatment during follow-up.

**Conclusions:** A single, comprehensive surgery session was able to significantly lower IOP, reduce GMS and control pain.

## Introduction

An estimated 75,000 to 113,000 individuals are affected by neovascular glaucoma (NVG) in the European Union ^1^. A variety of ocular and systemic conditions can cause retinal hypoxia that eventually leads to neovascularization of the iris (NVI) and the angle (NVA). NVG carries a poor prognosis for both ocular and general health: at five years, the failure rate of glaucoma drainage implants to control intraocular pressure (IOP) are near 80% ^2^ and of cyclophotocoagulation near 65%, respectively; remarkably, patient survival itself is reduced by 40% at five years ^3^.

Neovascularization is the formation of abnormal blood vessels in an abnormal location. In NVG, vessels and fibrovascular membranes block aqueous humor outflow in the angle of the anterior chamber. The most common predisposing ocular conditions for NVG are retinal ischemia caused by central retinal vein occlusion (CRVO), proliferative diabetic retinopathy (PDR), central retinal artery occlusion (CRAO), and ocular ischemic syndrome ^1^. Carotid artery obstructive disease and fistulas are additional, extraocular vascular causes for retinal ischemia ^1^. Early recognition and therapy are imperative to prevent aggressive evolution with severe vision loss and intractable pain. In the past, treatment steps have been mostly applied sequentially. These include IOP lowering (topical and systemic glaucoma medications, glaucoma drainage implants, cyclodestruction) ^4–8^, anti-inflammatory treatment (topical or intraocular steroids) as well as treatment of retinal ischemia (panretinal photocoagulation (PRP) ^9,10^, and vascular endothelial growth factor (VEGF) inhibitors ^11–13^) as an adjuvant therapy. Other studies indicate that vitrectomy with PRP and silicone oil tamponade may reduce IOP in eyes with NVG ^14,15^. Although NVG can quickly lead to a blind and painful eye that requires enucleation ^16^, there is no general consensus on how to best initiate treatment.

We evaluated the safety and efficacy of an integrative combined surgical approach for evolving NVG that combined pars plana vitrectomy, near-confluent full-scatter panretinal photocoagulation, intravitreal bevacizumab, and transscleral cyclophotocoagulation in one operation. Phakic eyes underwent concomitant cataract surgery. We hypothesized that this approach would prevent or slow NVG progression when applied at first sight of neovascularization and IOP elevation > 21 mmHg.

## Methods

### Study design

Our study was reviewed and approved by the Ethics Committee of the University of Würzburg, and performed in accordance with the ethical standards set forth in the 1964 Declaration of Helsinki and the Health Insurance Portability and Accountability Act.

This consecutive interventional case series included patients who met the following inclusion criteria: 1) NVI or NVA, 2) IOP > 21 mmHg, 3) visual acuity of at least light perception and 4) above or equal to 18 years of age. All patients were treated at the University Eye Hospital in Würzburg, Germany, between October 2014 and December 2018. We recorded best-corrected visual acuity (BCVA, logMAR), intraocular pressure (IOP, mmHg), glaucoma medication score (GMS) ^17^, visual analog pain scale (VAPS, 0-10) at baseline and at visits at 1, 3, 6 and 12 months.

Hypotony was defined as IOP ≤ 5 mmHg with hypotonous maculopathy, choroidal folds, or optic neuropathy ^18^. Phthisis bulbi ^19^ was defined as IOP ≤ 5 mmHg in a shrunken eye with worse than hand motion vision with or without pain containing atrophic and disorganized intraocular structures. Success was defined as lOP ≤ 21 mmHg or IOP reduction ≥ 30% from baseline, with or without glaucoma medication and without vision loss ^20^.

Eyes with no light perception and patients with a history of glaucoma other than NVG were excluded. All patients underwent decimal visual acuity testing which was converted to a logMAR scale. Counting finger (CF), hand movements (HM), light perception (LP), and no light perception (NLP) were converted into logMAR units 1.9, 2.3, 2.7, and 3.0, respectively ^21–23^.

### Surgical technique

All eyes underwent standard 3-port 23 gauge pars plana vitrectomy with detachment of the posterior vitreous if not already present, near-confluent full-scatter panretinal photocoagulation applied under indentation in all four quadrants from the vascular arcades to the ora serrata, intravitreal 0,1 mL of bevacizumab (Avastin® 25 mg/1 mL, Roche Pharma, Switzerland), transscleral cyclophotocoagulation (810 nm diode laser, 360 degrees treatment to pop threshold with 20 spots and leaving out 3 and 9 o’clock), and air tamponade. Phakic eyes underwent concomitant cataract surgery using a bag-in-the-lens IOL (type 89F; Morcher, Stuttgart, Germany) to reduce the incidence of posterior capsular opacification and posterior synechiae ^24,25^. The operation was carried out either with a retrobulbar block or general anesthesia.

### Retreatment

At follow-up, all eyes with elevated IOP were treated following an escalation scheme. First, glaucoma medications were increased to what was maximally tolerated. Eyes were then treated with transscleral cyclophotocoagulation (810 nm diode laser, 360 degrees treatment to pop threshold with 20 spots). Eyes that failed to respond to cyclophotocoagulation with a significant IOP reduction and had retained ambulatory visual acuity underwent tube shunt surgery. Repeated vitrectomy including fill-in panretinal photocoagulation, transscleral cyclophotocoagulation and intravitreal bevacizumab was applied in eyes with elevated IOP and dense vitreous hemorrhage. Fill-in panretinal photocoagulation was applied in these eyes when panretinal photocoagulation had not been completed in the first vitrectomy due to extensive intraretinal hemorrhage in ischemic retinal disease. Further intravitreal injections of VEGF inhibitors were only applied for clinically significant non-ischemic macular edema and BCVA ≥1.1 logMAR.

### Statistical Methods

Data analysis was performed using Statistica 13.1 (Tulsa, Oklahoma, United States). Categorical variables were described by the frequency of observations. Continuous variables were described as mean with standard deviation (SD) or median with range (minimum – maximum). Friedman test and Wilcoxon signed-rank test were used to compare data measured on an ordinal scale and continuous variables with non-normal distribution. Evaluation of data normality was performed using the Shapiro-Wilk test. Welch’s t-test for unequal variances was used for IOP between pain versus no-pain group comparison. Kaplan–Meier curve and log-rank test were used for success analysis. P-values <0.05 were considered significant.

## Results

83 eyes of 83 patients (53 male, 30 female, mean 74.6±11.6 years) were included of which 1 month, 3 months, 6 months and one year follow up completed 81, 71, 62 and 53 eyes, respectively. Conditions that lead to NVG included CRVO (n=41), PDR (n=25), CRAO (n=10), and ocular ischemic syndrome (n=7).

Mean logMAR BCVA was 2.0±0.7 at baseline and 1.8±0.8 at twelve months (p=0.47, **Fig.1**). Three eyes worsened from LP to NLP at one week, six months, and twelve months, respectively. At baseline, 32.5% (27/83) of patients and at 12 months 47.2% (25/53) of patients had an ambulatory visual acuity (≥ logMAR 1.7), respectively. Only two patients presented at baseline with IOP<30 mmHg.

**Fig. 1.**
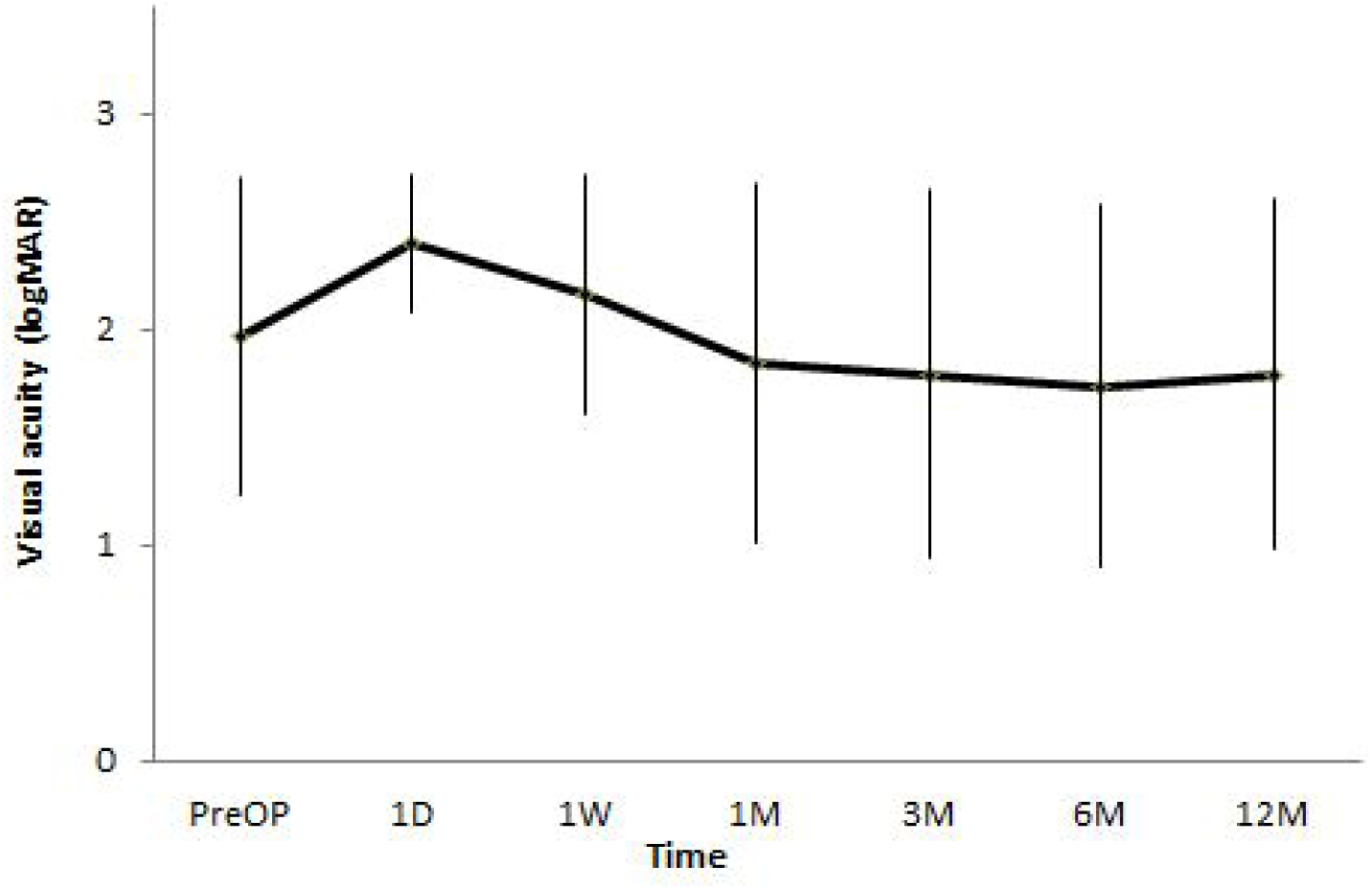
Diagram of visual acuity during follow-up (mean ± SD) (p=0.47).

IOP decreased significantly from the baseline of 46.0±10.3 mmHg at one month (18.8±9.9 mmHg), three months (17.2±9.8 mmHg), six months (14.7±8.1 mmHg), and twelve months (14.2±8.9 mmHg; p<0.001; **Fig. 2**). At one-year follow-up, 90.6% (n=48) of patients had an IOP ≤ 21 mmHg. IOP ≤ 5 mmHg was found in 11.3% (n=6) and tolerated without complications.

**Fig. 2.**
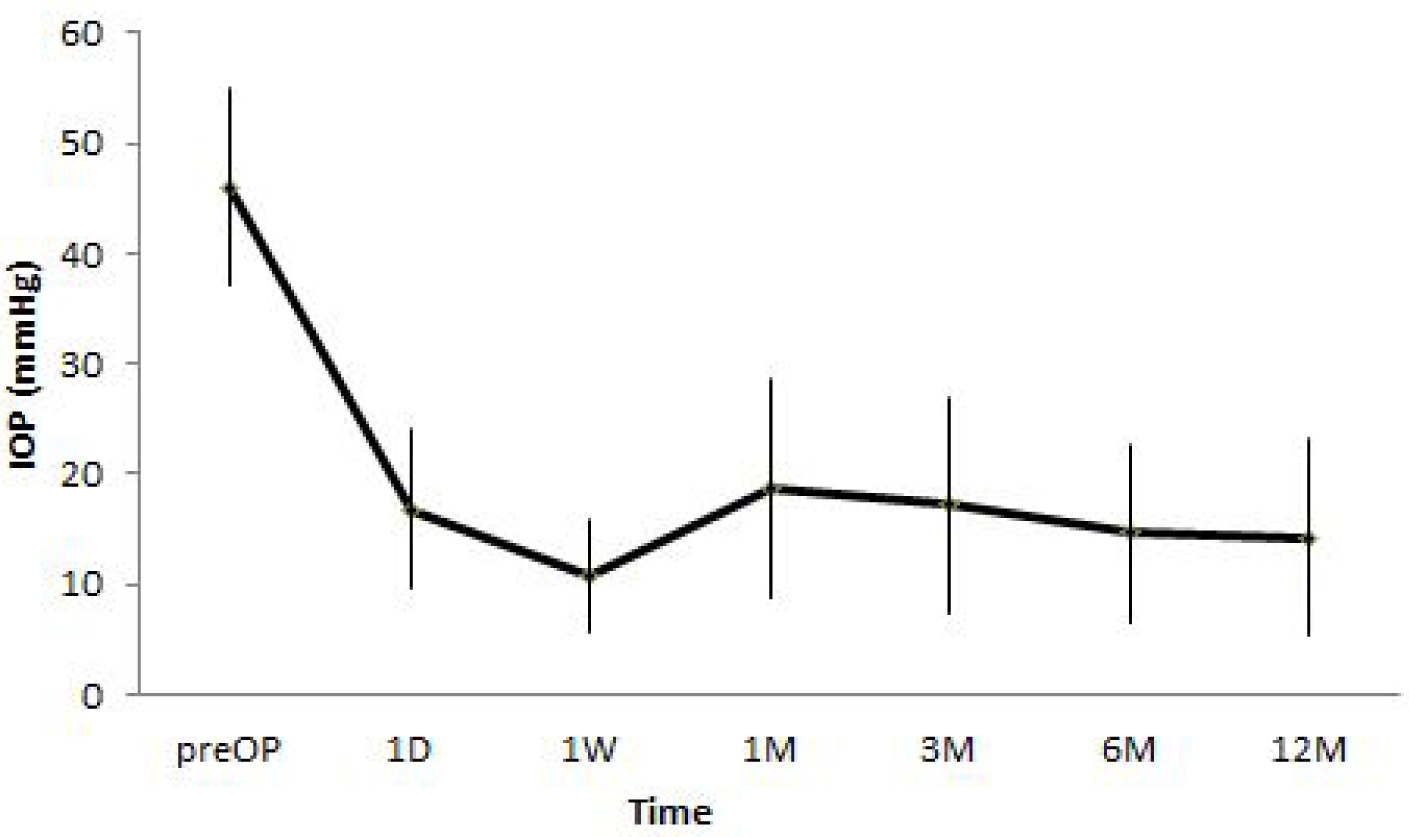
Diagram of IOP during follow-up (mean ± SD) (p<0.001).

GMS decreased from 4.8±2.5 medications at baseline to 1.8±1.8 (p<0.001; **Fig. 3**) at twelve months. While 32.5% (n=27) of patients complained of ocular pain at baseline (VAPS: 6.3±1.6) all patients were without pain at all follow-up visits. Patients with pain had a significantly higher baseline IOP of 49.2±7.9 mmHg compared to patients without pain 44.5±11.1 mmHg (p=0.027).

**Fig. 3.**
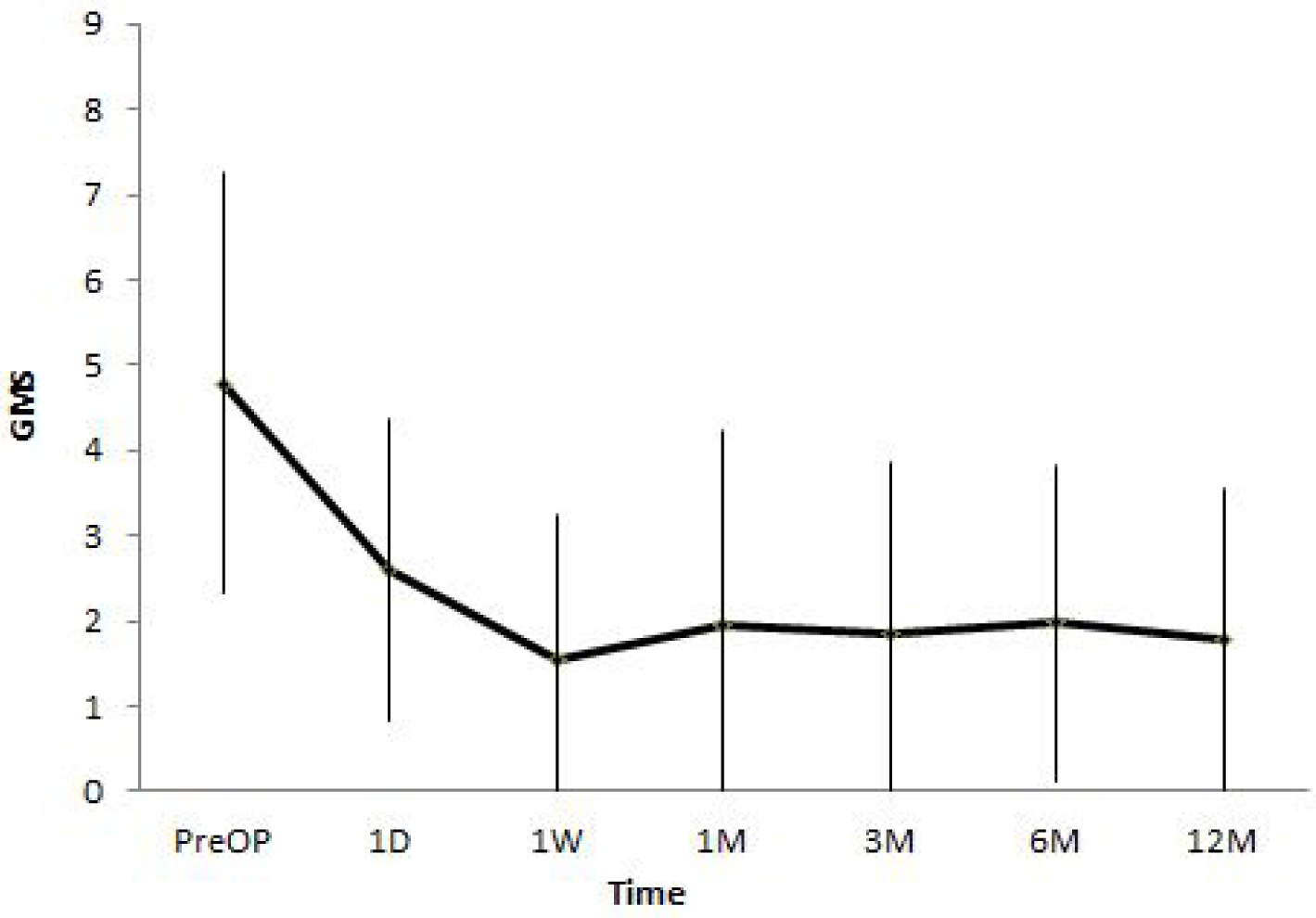
Diagram of GMS during follow-up (mean ± SD) (p<0.001).

Early postoperative complications (day 1–4 weeks) included intraocular fibrin (56/83), hyphema (18/83), choroidal detachment (14/83), and corneal erosion (12/83) **(Table 1)**. Late postoperative complications (> 1 months) included retinal detachment (3/83) and painless phthisis bulbi (4/83).

**Table 1.** Early and late complications.

|  | early complications |  | late complications |  |
| --- | --- | --- | --- | --- |
|  | n | % | n | % |
| hyphema | 18 | 21.7 | 0 | 0.0 |
| fibrinous reaction | 56 | 67.5 | 0 | 0.0 |
| hypopyon | 0 | 0.0 | 0 | 0.0 |
| corneal erosion | 12 | 14.5 | 0 | 0.0 |
| choroidal detachment | 14 | 16.9 | 0 | 0.0 |
| retinal detachment | 1 | 1.2 | 3 | 3.6 |
| phthisis | 0 | 0.0 | 4 | 4.8 |
| hypotony | 0 | 0.0 | 3 | 3.6 |

Hypotony: IOP ≤ 5 mmHg with hypotonous maculopathy, choroidal folds, or optic neuropathy <sup>18</sup>; phthisis bulbi: IOP ≤ 5 mmHg in a shrunken eye with little or no vision with or without pain containing atrophic and disorganized intraocular structures <sup>19</sup>.

During one-year follow-up, retreatment was applied in 27/53 eyes. 44.4% (12/27) underwent one additional cyclodestruction. Other re-treatments were vitrectomy (8/27), Baerveldt glaucoma drainage (5/27), trabeculectomy (1/27), and filtering bleb needling (1/27). 7.5% (4/53) patients with a mean BCVA 1.1±0.3 logMAR received further anti-VEGF injections (4.0±0.8 injections).

27.7% (23/83) patients were lost to follow-up and 8.4% (7/83) patients died during the one-year follow-up.

Kaplan-Meier analysis shows a probability of success of 65% at one year follow-up **(Fig. 4)**.

**Fig. 4.**
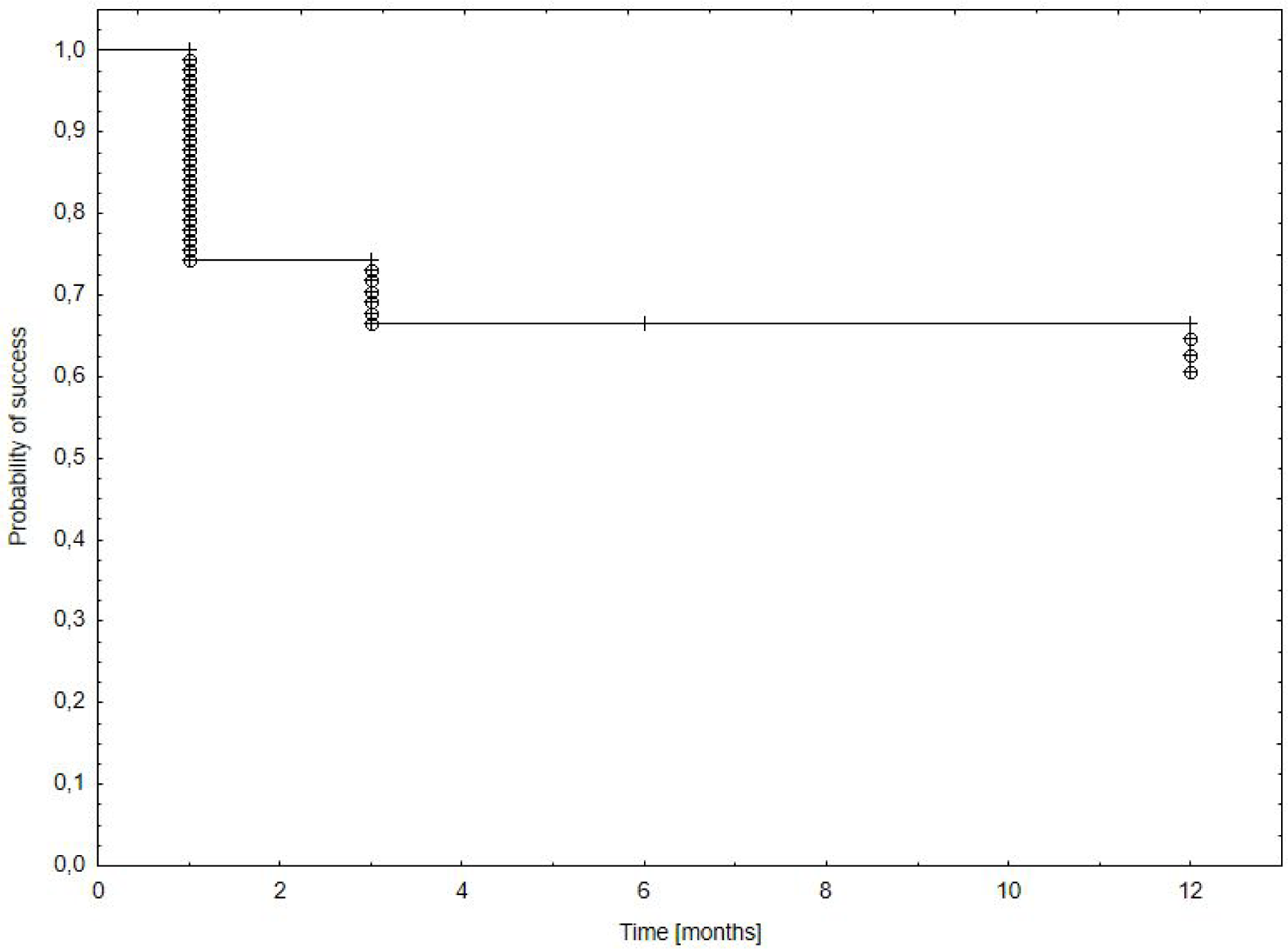
Kaplan-Meier univariate estimates of the probability of success: lOP ≤ 21 mmHg or IOP reduction ≥ 30% from baseline, with or without glaucoma medication, without vision loss.

## Discussion

By 1871, NVG was known as “glaucoma haemorrhagicum et apoplecticum” and feared as a consequence of ischemia that quickly led to enucleation due to “hefty ciliary neuralgia” ^26^. In 1963, using improved equipment, Weiss et al. found that emerging neovascularization and fibrovascular membranes of the iris and the angle could be observed well before the onset of advanced NVG ^27^ hinting at a window to initiate treatment. Today, the ability to detect neovascularization early is complemented by better interventions that address both the underlying pathology and the elevated IOP. However, these interventions need to be implemented urgently because NVG continues to have a rapid evolution and a poor prognosis for both the eye and the patient ^3^.

Addressing this need, our results show that the combined approach of lens extraction with bag-in-the-lens IOL, pars plana vitrectomy, near-confluent full-scatter panretinal photocoagulation between the vascular arcades and the ora serrata, intravitreal bevacizumab, and transscleral cyclophotocoagulation is a safe and effective treatment that can lower mean IOP from about 46 to near 14 mmHg by post-op day one, reduce the number of glaucoma medications and control pain. Retinal ischemia is the primary cause of neovascular glaucoma ^1,28^. Accordingly, PRP is the standard of care to reduce posterior pole oxygen demand and angiogenic drive while vitrectomy is performed to increase the partial pressure of vitreous oxygen ^28–31^. We performed lens extraction, pars plana vitrectomy, and endoscopic PRP because of significant media opacities and because endoscopic laser through the pars-plana approach facilitates the delivery of 360° near-confluent peripheral retinal laser treatment out to the ora serrata. In our clinical experience, further progression of NVG with elevated IOP can be maintained by even relatively small areas of untreated ischemic retina. Therefore, we took great care to apply 360° PRP to near-confluence up to the ora serrata. Such extensive treatment would be more difficult to accomplish using standard externally delivered PRP.

In the healthy eye, the vitreous body and the iris-lens diaphragm form a relative diffusion barrier that maintains a higher oxygen partial pressure in the anterior chamber compared to the vitreous overlying the posterior pole. Concurrently, it reduces the diffusion of angiogenic mediators. Recreating a relative diffusion barrier after vitrectomy ^30^ is beneficial and reduces the occurrence of NVI ^32^. A barrier can be achieved with silicone oil ^33^ that lessens the incidence of neovascular glaucoma ^33,34^. Following this concept, Bartz-Schmidt et al. treated 32 NVG patients with pars plana vitrectomy, retinal and ciliary body photocoagulation, as well as silicone oil tamponade because eyes were left aphakic ^14^. This approach controlled IOP in 72% of patients for at least one year. In our study, all eyes were pseudophakic and without silicone oil at the conclusion of the surgery, thereby negating the need for silicone oil as a diffusion barrier. We observed IOP control in 79,2%, more eyes than reported by Bartz-Schmidt et al. ^14^.

Our success rate of 65% defined as IOP <22 mmHg, with or without glaucoma medication and without vision loss after one year follow-up is similar to success rates reported for glaucoma drainage devices which range from 62% to 66.7% ^8,35–37^. One study reported 73% success rate in 38 eyes treated with glaucoma drainage device and, in contrast to other studies ^8,35–37^, with very few postoperative complications ^2^. The integrative surgical approach presented here avoids tube-specific complications (e.g. tube exposure, retraction, corneal touch, obstruction) that can range from 13% to 26% in NVG over five years ^2,8,35–37^.

Our approach delivers both retina and glaucoma treatment in a single surgical session and reduces the burden on the patient and health care system by simplifying postoperative care and follow up. It is worth noting that an IOP reduction could also be achieved without cyclodestruction applying only pars plana vitrectomy, lensectomy with a preserved anterior capsule, and panretinal endophotocoagulation. However, only 13 eyes were included and the mean preoperative IOP of 29 mmHg was lower than in our study ^15^. By contrast, treatment with anti-VEGF agents was insufficient as a primary therapy ^12,38^, with failure rates up to 88% ^38^. Anti-VEGF agents may be best used as an adjuvant treatment ^13^. Consistent with our findings, pars plana vitrectomy, endoscopic peripheral panretinal photocoagulation, and endocyclophotocoagulation (ECP) also led to an IOP reduction and seemed to be more effective compared to panretinal photocoagulation, intravitreal bevacizumab, pars plana vitrectomy, and filtration surgery or Ahmed valve placement, but in this study phthisis bulbi occurred in 7.4% of cases ^39^.

Unsurprisingly, visual function remained poor in most eyes in our study. Although 67% of eyes were inflamed postoperatively, inflammation was transient and painless in all eyes. The observed high mortality rate is a reminder that any treatment strategy of NVG should ideally require as few surgical interventions and hospital visits as possible.

Our study has several limitations. Because the integrative surgical approach described here was the primary practice pattern, there was no control group. This limited us to an intragroup comparison of before versus after treatment data. As a retrospective study, in addition to informing on parameters and design of future prospective studies, it can only help to formulate, but not answer, hypotheses about associations between treatment and outcomes.

In conclusion, this study shows that NVG can be controlled by an integrative surgical approach delivered in a single session that combines cataract removal, pars plana vitrectomy, near-confluent full-scatter panretinal photocoagulation, intravitreal bevacizumab, and transscleral cyclophotocoagulation. This approach simplifies care and addresses multiple ophthalmic and patient-related challenges.

## Data Availability

The datasets generated during and/or analysed during the current study are available from the corresponding author on reasonable request.

## Notes

### Competing Interest Statement

The authors have declared no competing interest.

### Clinical Trial

We conducted a retrospective study at our University Hospital and did not register this study.

### Funding Statement

None.

